# Ranking Radiology Residency Programs: What Medical Students Think is Important Versus What Residents Say is Important

**DOI:** 10.1101/2020.06.29.20143008

**Authors:** Florentino Saenz Rios, Shadan Alwan, Quan Nguyen

## Abstract

**Background:** Matching to a residency is a great success, but it is also a significant change in the lives of many graduating medical students. It is expected that during this time, many priorities may change, especially after participation in a residency program where residents are further exposed to the dynamics of the program.

**Purpose:** There are no current studies that attempt to determine what factors residents prefer after the match.

**Methods:** This study was an anonymous study conducted through an online survey. The survey asked two open-ended questions asking the survey taker to list four factors in order of importance that they considered important in a program, one as an MS4 and one as a resident. Results were compiled, tallied, and categorized to find common themes between the applicant’s preferences through descriptive statistics.

**Results:** Of the 24 surveys sent out, a total of 15 applicants responded for a response rate of 62.5%. The most common preferences among both MS4s were “Work Environment” and “Location,” however, factors like “Education” and “Faculty: Resident Ratio” was seen as increasing in importance among residents.

**Conclusion:** While an increase was seen in some factors (Education, Faculty:Residency Ratio, and Program Size), the general trend of preferences going from MS4 to a resident was more spread-out in the distribution of what residents considered essential factors. Education involves both formal didactics and readout of dictated exams. Higher Faculty:Residency affects education because more faculty decreases the clinical workload of each faculty and leaves the faculty more time for teaching. Larger program size decreases the amount of call burdern per resident.

## Introduction

The NRMP match process is a considerable change in the lives of many graduating medical students; the match is a time of many unknowns with varying factors influencing how applicants rank specific programs within the Diagnostic Radiology match. Many studies have observed what factors these applicants prefer, attempting to discern trends that residency programs could use in order to improve their respective programs, increasing the quality of matched applicants in the process.^[1, 3–6, 7, 9–12, 14]^ However, many applying medical students are inexperienced when it comes to the dynamics of participating in a residency program with many unknowns being left until after the beginning of residency. It is possible that once an applicant has gained experience as a resident that a shift in what factors they value in a program may be seen.^[10, 14,16]^ Residency programs want to attract the most applicants with the best qualifications and the most exceptional fit for the program. These programs must be able to not only understand but develop a model that coincides with what modern Diagnostic Radiology requires to create a fulfilling career.

This study is novel in that only residency program applicants are surveyed in order to determine what factors residency programs should consider when preparing for the match process in order to achieve the resident-program fit both during and after the NRMP match. This type of data is vital in assuring that the program’s residents have a voice in the development of their program as well as exposing future potential Diagnostic Radiology applicants to residency program ranking factors they had not previously considered. ^[7, 11–14, 16, 18]^ In addition to this, by exposing these factors, it may potentially decrease the rate of future burnout associated with applicants going to specific programs for reasons that are not compatible with their idea of job satisfaction. ^[2–4, 7, 9, 11, 13, 17]^ A pilot study such as this one is important in establishing the potential for further trends to be seen among Diagnostic Radiology residents, trends that when further explored could potentially help program directors modify their program’s curriculum to increase the overall quality of resident education with minimal wasted resources.

## Methods

This study was designed as an anonymous cross-sectional study conducted through an online survey hosted by the survey site Survey Monkey™ (Palo Alto, Ca). Emails containing links to the survey were sent out to Diagnostic Radiology residents at the University of Texas Medical Branch (UTMB) within different years of the program. This study received IRB approval from the UTMB IRB as well as an exemption from requiring informed consent due to the anonymous design of the study. No compensation was offered to candidates for their participation in the survey.

### Survey Enrollment

This study aims to differentiate between what factors Diagnostic Radiology applicants consider essential in a program both right after they matched as a fourth-year medical student (MS4) and after at minimum a year of practice at the Diagnostic Radiology residency program at UTMB (Galveston, TX). Resident emails were obtained from the UTMB email directory that is available to all \ within the UTMB healthcare system. In total, 24 emails were gathered and promptly sent out with the open-ended survey on March 16^th^, 2019. In the email containing the survey link, residents were made aware that the link would be available for two weeks, closing on March 30^th^, 2019. UTMB residents were informed via email on the anonymity of the survey, the purpose of the survey, and the availability of the option to opt-in or out by responding or not responding to the survey.

### Development of the Survey

The survey protocol used in this study was developed through a review of recent literature in addition to discussion among the investigating authors, and input provided by the UTMB Department of Radiology faculty. To ensure anonymity between participating residents in addition to allowing residents to have the full ability to express what they believe is essential in a residency program, it was decided that the online survey website Survey Monkey would be used to gather responses. The survey design was developed to include two mandatory open-ended questions in order to maximize the number of responses in addition to eliminating any bias from predetermined answer choices. Each question asked was designed to elicit the top four factors the resident considered most important in a Diagnostic Radiology residency program both after their match as a fourth-year medical student and after at least a year of practice at a Diagnostic Radiology program. Participating residents who chose to take the survey were instructed to “Describe the top four factors (from most to least important) you considered when creating your residency program rank list?” and to “Describe the top four factors (from most to least important) you consider in a Diagnostic Radiology after practicing at a residency program?” Once the survey was completed, the answers to the open-ended question were compiled, grouped into categories, and tallied amid discussion by the investigating authors.

### Disbursement of the Survey

Emails to UTMB residents containing information on the anonymity of the study, the purpose of the study, and the survey link were sent through the blind “carbon copy” feature in order to ensure anonymity between residents participating. Initial emails containing this information was sent to each potential participant on March 16^th^, 2019, with a notice that the survey would only be open for two weeks until March 30^th^, 2019. Only surveys completed before closure of the survey on March 30^th^, 2019, were considered during the data analysis.

## Results

A total of 24 emails (n = 24) were gathered from the UTMB system email directory with 24 email invitations to participate in the study being sent out to residents at different years of training. Of these 24, 15 UTMB residents responded to and completed the survey before the set date, giving the survey questions a response rate of 62.5%. Responses to the 15 completed surveys were compiled, grouped into categories, and tallied after discussion by the investigating authors. Grouping of resident’s responses found the standard answers to fall under the factors “Location,” “Reputation,” “Work Environment,” “Education,” “Work-Life Balance,” “Fellowships,” “Board Pass Rate,” “Moonlighting,” “Program Size,” “Faculty:Resident Ratio,” “Family & Friends,” “Case Volume/Diversity,” “Research,” and “Other.” The factor “Other” was reserved for answers that did not fit within any other of the other factors decided on. Common examples that were categorized under “Other” include benefits/book and travel funding, ancillary opportunities, visa sponsorship, gut feeling, organization/willingness to change, and program diversity. Due to the themes in “Other” not being common to other responses, the “Other” factor will not be discussed in this analysis.

### Top Factors Considered a 1^st^ Preference Pre and Post Residency

The top factor that the then MS4s considered prior to joining UTMB Diagnostic Radiology Program, as per Figure 1, is “Location” at 46.7%, with “Work Environment,” the only other factor receiving as many mentions for most crucial factor.The only other factors being ranked are “Friends and Family,” “Fellowships,” and “Board Pass Rate,” all being tied for MS4s most important residency factor at 6.7%, albeit mentioned less often than “Location” and “Work Environment.” The two factors, “Location” and “Work Environment,” remained to be factors significant to residents, as per Figure 2, both being tied at 26.7% for the most essential factor.

**Fig 1.**
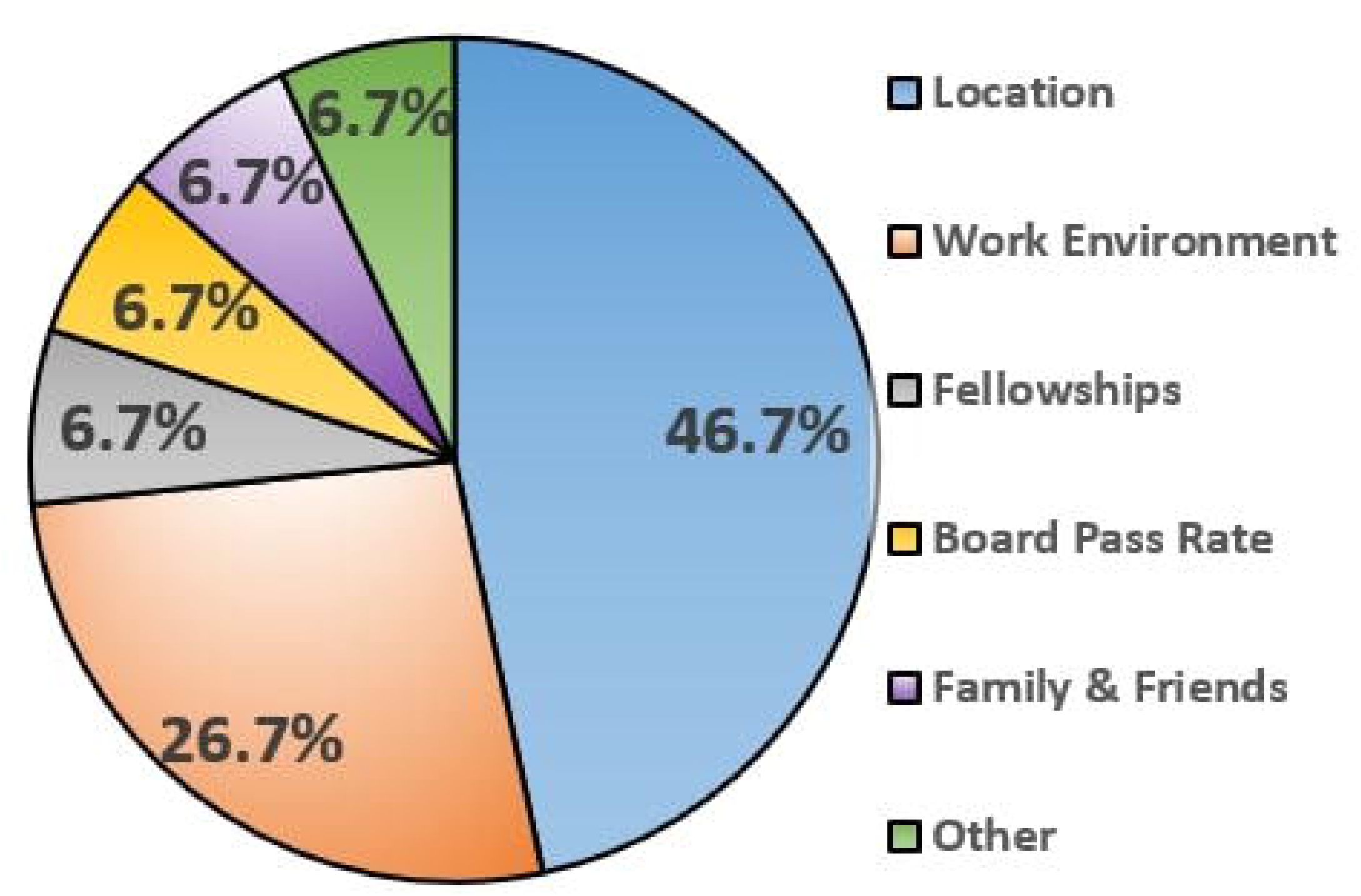

**Fig 2.**
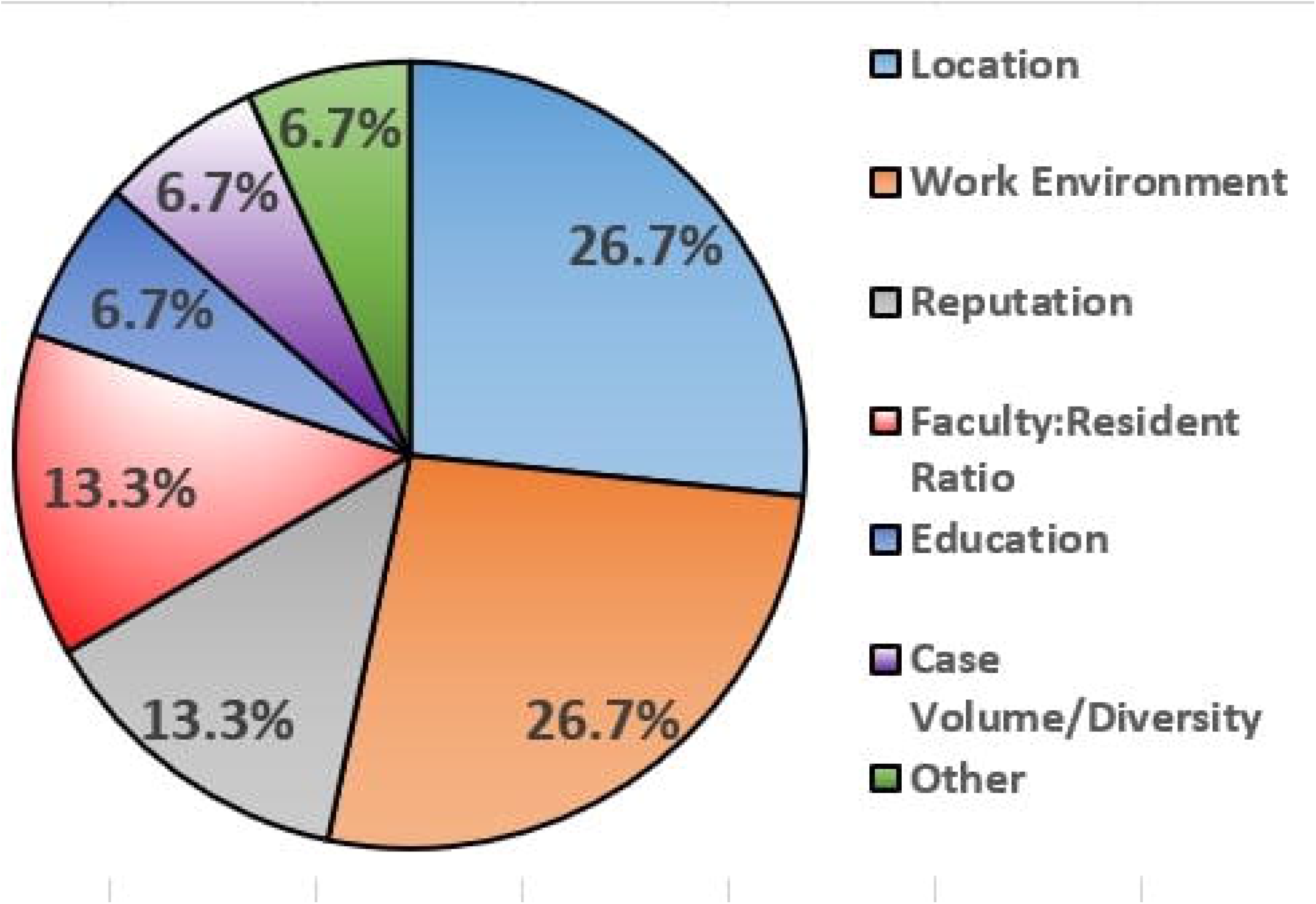

### Top Factors Considered A 2^nd^ Preference Pre and Post Residency

A similar shift from MS4 to resident can be seen in the 2^nd^ most preferred factors with “Work Environment” being the second most 2^nd^ preferred factor for MS4s at 40% now followed by “Reputation” at 26.7%, as per Table 1. “Location” trails both of these at 20% with the only other factor being mentioned as 2^nd^ most preferred factor being “Education” at 6.7%. The 2^nd^ most preferred factors for residents are “Work Environment” at 33.3%, followed by “Education” at 26.7%, and “Reputation” at 33.3%.

**Table 1:**
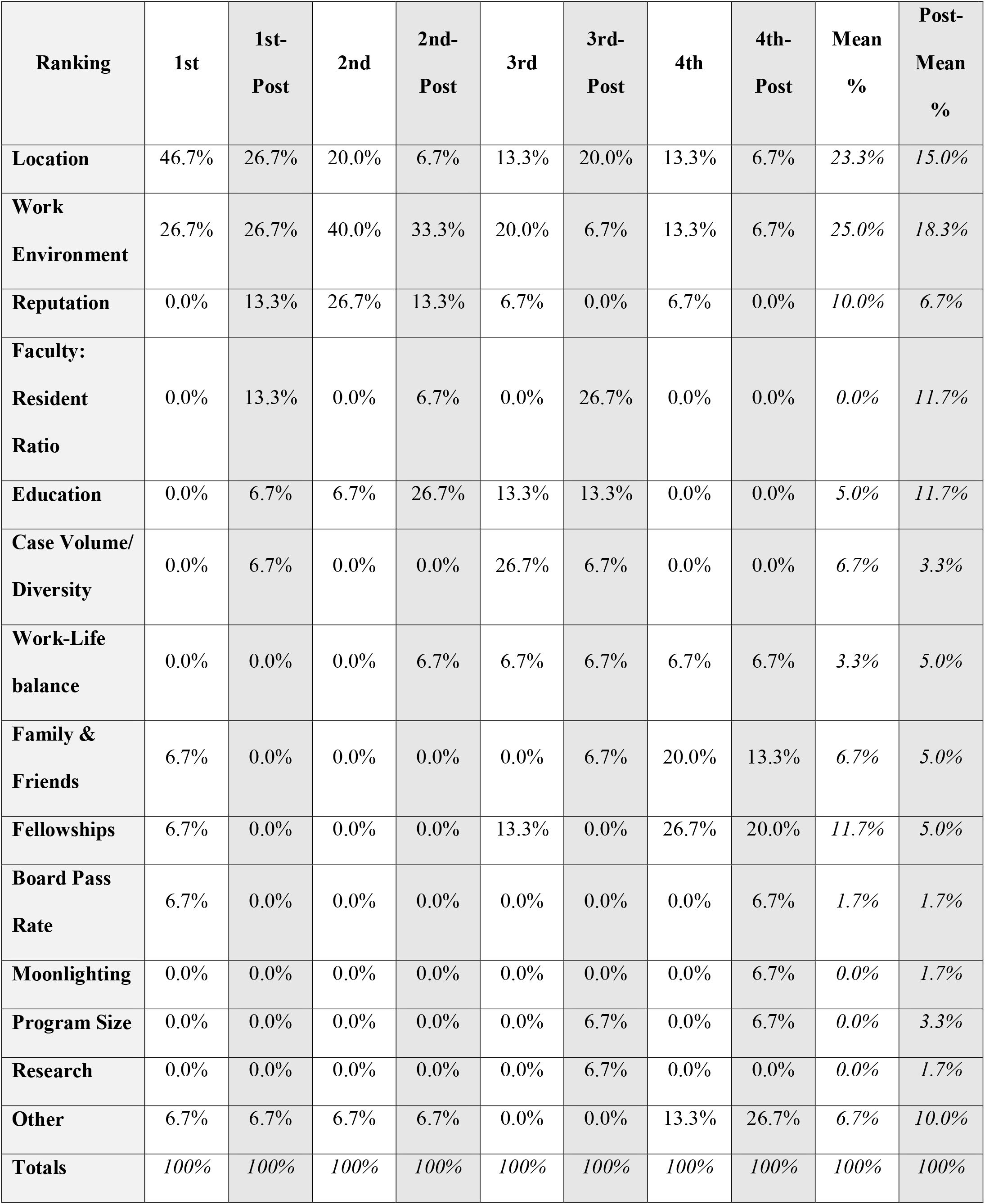
Shifts in Responses to the Importance of Factors in a Residency Program between MS4s and DR Residents.

### Factors Considered A 3^rd^ Preference Pre and Post Residency

Looking at Table 1, it is seen that the factor that was most often mentioned as the residents’ 3^rd^ top factor prior to their experience at a residency program was the factor “Case Volume/Diversity” at 26.7% with the factor “Work Environment” trailing at 20%. After “Work Environment,” the factors “Location,” “Education,” and “Fellowships” were all mentioned as 13.3% of residents’ 3^rd^ top factor. The factor that most residents believed was 3^rd^ most important was the “Faculty: Resident Ratio” at 26.7%, which was closely followed by “Location” at 20%. The factor “Education” was mentioned as the 3^rd^ preference by 13.3% of respondents. Interestingly, the factors “Work Environment,” “Case Volume/Diversity,” “Work-Life Balance,” “Family & Friends,” “Program Size,” and “Research” all tied together with 6.7% of respondents choosing these as their 3^rd^ most important factor they considered as an MS4.

### Top Factors Considered A 4^th^ Preference Pre and Post Residency

The residents’ fourth most important factor when they were MS4s was the factor “Fellowships,” placing highest at 26.7% with the factor “Family & Friends” continuing to trail it now at 20%. Following these factors are “Location” and “Work Environment,” tied with 13.3% of residents believing this is the 4^th^ most important factor in a residency program. The fourth most considered factor for the residents was the factor “Fellowships” at 20%, which was followed by the factor “Family & Friends” at 13.3%. After these two, there is a six-way tie between “Location,” “Work Environment,” “Work-Life Balance,” “Board Pass Rate,” “Moonlighting,” and “Program Size” at 6.7%.

## Discussion

Determining what factors residents value in a residency program is an essential piece of information necessary for a program to align itself with the interest of its residents further, allowing both the program and its residents to grow and prosper. While each program has different dynamics that may make generalizability difficult, this study aims to highlight some interesting points that may not be commonly discussed or even thought about. Some of these points could allow program directors and program committees to modify their program’s growth plan to fit what their residents value the most more accurately. The findings of this study help us to better understand “how do a medical student’s priorities change after matriculating into a program?” and “what are the most important factors to change to meet the needs of the residents?” It is important to note that the survey was disbursed to residents with different years of training at the program, which may be an indicator of why results seen from the residents after their residency program exposure seems to be so dispersed. However, this may be a sign that after acceptance into a residency program, residents were able to experience all of the dynamics of not only the radiologist’s career but of the residency itself. These changes may have played a part in the shift in priorities that can be seen when comparing the MS4 results to the resident results. While the sample size of this study (n = 15) was a limiting factor, we believe the overall design, communication, and timing of the survey’s disbursement was able to convince 15 participants to respond to the survey, giving the survey a response rate of approximately 62.5%.

### Factors Most Commonly Preferred As an MS4 vs. Resident

Once tallied the results show that the factors that were thought to be important in choosing a residency by the MS4s were seen to be both “Location” at 46.7% and “Work Environment” at 26.7%, with “Family & Friends,” “Fellowships” and “Board Pass Rates” at 6.7% as shown in Figure 1. A substantial shift in priorities can be seen when comparing these numbers to the post-residency participation numbers where the factor “Location” ended up decreasing from 46.7% to 26.7%. While “Location” received a decrease in preference among residents, the factor “Work Environment” neither had a loss nor gained in preference 26.7%. It is worth noting that other than these two factors the other factors such as “Reputation,” “Faculty: Resident Ratio,” “Education,” and “Case Volume/Density” all increase from 0% in the MS4 preferences to 13.3%, 13.3%, 6.7%, and 6.7% respectively. In addition, a decrease of 6.7% was seen in the factors “Family & Friends,” “Board Pass Rate,” and “Fellowships” among residents, likely hinting at there being an underlying change in life or career goals that a resident may develop as they mature throughout their career. It is also interesting to note that while some of the mentioned factors are not capable of easily being changed by the program such as the program’s location or the surrounding family/friends, there are modifiable factors such as “Work Environment,” “Education,” and “Faculty: Resident Ratio” that programs can work on. By understanding these preferences, residency programs can more effectively grow towards achieving goals that would attract potential fellows while also keeping their residents satisfied.

### Factors Most Commonly Mentioned As an MS4 vs. Resident

While it is essential to consider the factor that most residents considered their highest priority, it is also essential to look for any trends in the data itself. Table 1’s “mean percentage” and “post-mean percentage” columns are the aggregate averages of the number of times each factor was listed as a primary, secondary, tertiary, or quaternary preference. With that in mind, the means of each factor was found in order to discern how often a preference was mentioned as one of the top four factors, allowing any patterns in terms of significant shifts in preference during the shift from MS4 to resident. Something to take note of is that while a noticeable shift did happen in the transition from MS4 to resident, two factors stayed relatively constant as seen when comparing these means to the raw percentages in Figure 3 and Figure 4. These two factors were “Work Environment” (25% to 18.3%) and “Location” (23.3% to 15%), both of which received decreases in their percentages, remaining relatively constant. This consistency shows how important these are in defining a program’s identity. It is also worth noting that in the transition from MS4 to resident, factors such as “Faculty: Resident Ratio” and “Education” saw significant increases in overall mentions, going from 5% to 11.7 and from 0% to 11.7% respectively. At the same time, other factors such as “Fellowships” and “Case Volume/Diversity” had decreases going from 11.5% to 5.0% and 6.7% to 3.0% after this transition. The causes of these changes are not evident. However, the fact that they do change after the transition from MS4 to the resident can be indicative of a shift in priorities or a greater understanding of what is essential to a resident while practicing within the program.

**Fig 3.**
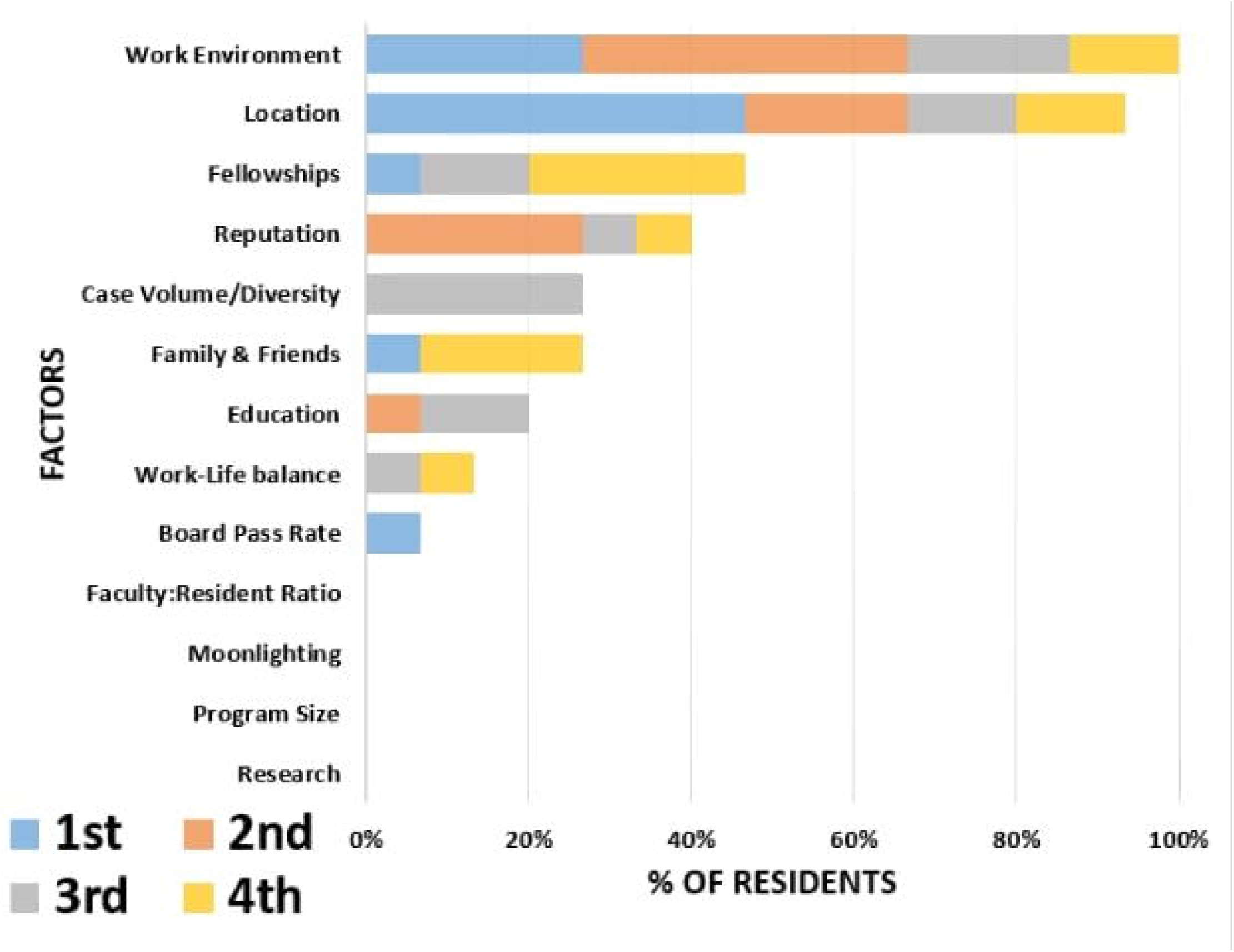

**Fig 4.**
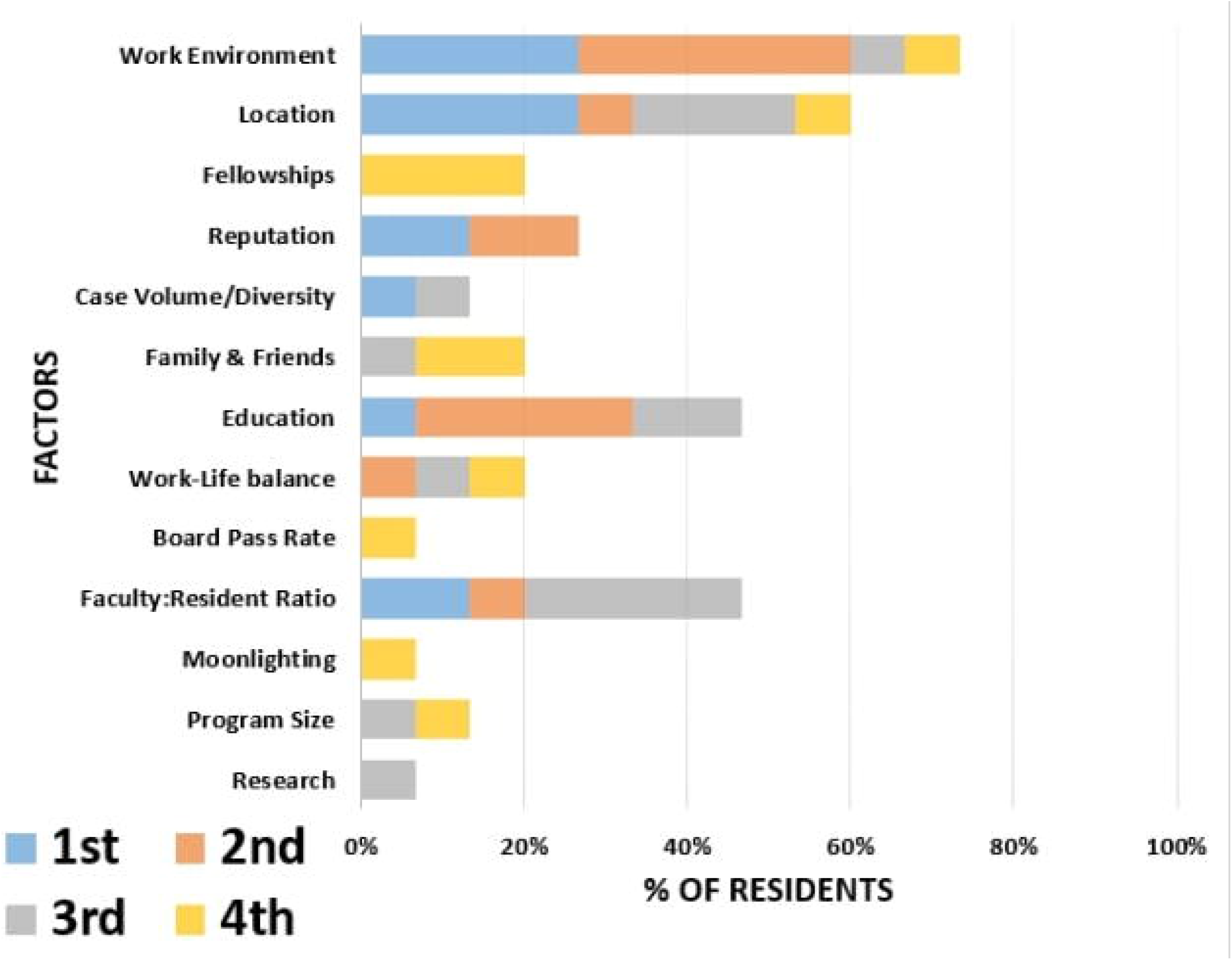

### Conclusions

Seeing as some of the most commonly mentioned factors for surveyed applicants were a program’s work environment in addition to its location, this study finds that it may be advisable for residency programs to invest in the development and discussion of a program with a collegial environment with a team player culture in order to effectively recruit and attract the residents with the best fit to their program. Due to the factor location being one that is not easily changed, program directors may find that discussing the program’s location with applicants to discern fit may help make sure that both the applicant and the program director are satisfied with the match. Regarding residents, it seems that the significant life change with beginning a career as a physician is a time where residents are beginning to explore available factors to them. Factors with low importance to MS4s such as “Moonlighting,” “Program Size,” and “Research” are seen as being at least somewhat important to residents, again likely identifying either a shift in their overall priorities or showing that the residents are beginning to understand the dynamics of working in a residency program. A similar effect can be seen when comparing the rates of the factors “Education” and “Faculty: Resident Ratio,” which were barely mentioned in the MS4 results but had gained significant increases in the results for the residents.

Interestingly, the preferences seen in the MS4 results are more concentrated into nine different factors, as seen in Figure 3, when compared to the more even distribution over 13 factors seen in Figure 4. These shifts once again show a higher degree of differing priorities upon joining the residency program. Future studies encompassing multiple institutions and larger sample sizes would need to be considered in order to ensure more definitive and generalizable results.

### Potential Benefits

This study helps identify factors that current Diagnostic Radiology residents consider necessary during their education at a residency program. Determining this type of data is essential in that it allows Diagnostic Radiology residency program directors to modify program curriculum and allocate program resources as efficiently as possible. In understanding what factors current residents find essential in the modern residency, program directors could increase the overall “fit” between the program and residents. Further potential benefits include an increase in ABR Core Exam board scores, higher fellowship match rates, and the potential for attracting higher-quality candidates into the residency program.

### Limitations

Limitations seen within this study include those seen in many similar survey-based studies. These limitations include intrinsic biases due to self-reporting data, misinterpretation of questions by responders, evasive answers, and misinterpretation of answers by the investigators. In addition, this study only included UTMB Diagnostic Radiology residents, which introduce limitations such as a reduced overall sample size, limited generalizability, and findings with possible bias affected by the UTMB Diagnostic Radiology residency curriculum. Other limitations include the lack of clear set definitions for what is considered “Reputation” or “Work-Life Balance,” which may have influenced the applicant’s answers when tallied and categorized. However, the most substantial improvement from this study would be increasing the overall sample size by including residents from other partnered Diagnostic Radiology residency programs, increasing the overall significance of the results found. Future investigations should be conducted to further refine and build upon the results of this study. In addition to including other Diagnostic Radiology residency programs, a comparison between what residents from different specialties consider essential in their program could be an appropriate continuation of this pilot.

## Data Availability

The authors confirm that the data supporting the findings of this study are available within the article [and/or] its supplementary materials.

## Citations

[1] Adeyekun AA. Residents’ Perception of Postgraduate Radiology Training in Nigeria. West African Journal of Medicine [Internet]. 2010 Jan 1 [cited 2020 Jan 16];29(5). Available from: https://www.ajol.info/index.php/wajm/article/view/68251

[2] Chetlen AL, Chan TL, Ballard DH, Frigini LA, Hildebrand A, Kim S, et al. Addressing Burnout in Radiologists. Academic Radiology. 2019 Apr 1;26(4):526–33.

[3] Darras KE, Worthington A, Russell D, Hou DJ, Forster BB, Hague CJ, et al. Implementation of a Longitudinal Introduction to Radiology Course During Internship Year Improves Diagnostic Radiology Residents’ Academic and Clinical Skills: A Canadian Experience. Academic Radiology. 2016 Jul 1;23(7):848–60.

[4] Donovan A. Radiology Resident Teaching Skills Improvement: Impact of a Resident Teacher Training Program. Academic Radiology. 2011 Apr 1;18(4):518–24.

[5] England E, Collins J, White RD, Seagull FJ, Deledda J. Radiology Report Turnaround Time: Effect on Resident Education. Academic Radiology. 2015 May 1;22(5):662–7.

[6] Forster BB, Whittall KP. Stress in Canadian radiology residency training: a national survey [Letter]. Canadian Association of Radiologists Journal; Montreal. 1998 Oct;49(5):349–50.

[7] Goldman D, Martin J, Bercu Z, Newsome J, Grimm L. Differential Motivations for Pursuing Interventional Radiology: Implications for Residency Recruitment. J Am Coll Radiol. 2019 Jan;16(1):82–8.

[8] Griffith B, Kadom N, Straus CM. Radiology Education in the 21st Century: Threats and Opportunities. Journal of the American College of Radiology. 2019 Oct 1;16(10):1482–7.

[9] Kelly AM, Cronin P, Dunnick NR. Junior Faculty Satisfaction in a Large Academic Radiology Department. Academic Radiology. 2007 Apr 1;14(4):445–54.

[10] Lam CZ, Nguyen HN, Ferguson EC. Radiology Resident’ Satisfaction With Their Training and Education in the United States: Effect of Program Directors, Teaching Faculty, and Other Factors on Program Success. American Journal of Roentgenology. 2016 Mar 9;206(5):907–16.

[11] Lourenco AP, Cronan JJ. Teaching and Working With Millennial Trainees: Impact on Radiological Education and Work Performance. Journal of the American College of Radiology. 2017 Jan;14(1):92–5.

[12] Matalon SA, Guenette JP, Smith SE, Uyeda JW, Chua AS, Gaviola GC, et al. Factors Influencing Choice of Radiology and Relationship to Resident Job Satisfaction. Curr Probl Diagn Radiol. 2019 Aug;48(4):333–41.

[13] McManus IC, Jonvik H, Richards P, Paice E. Vocation and avocation: Leisure activities correlate with professional engagement, but not burnout, in a cross-sectional survey of UK doctors. BMC Medicine. 2011;9.

[14] Millington S, Ball I, Seabrook JA, McCauley W. Attracting top residency candidates: a survey of important program attributes. CJEM: Canadian Journal of Emergency Medicine. 2005 Nov;7(6):411–4.

[15] Mok PS, Probyn L, Finlay K. Factors Influencing Radiology Residents’ Fellowship Training and Practice Preferences in Canada. Can Assoc Radiol J. 2016 May;67(2):99–104.

[16] Moriarity AK, Brown ML, Schultz LR. We Have Much in Common: The Similar Inter-generational Work Preferences and Career Satisfaction Among Practicing Radiologists. Journal of the American College of Radiology. 2014 Apr;11(4):362–8.

[17] Nazarian LN. Over-Regulating Radiology Residencies: The Unforeseen Costs. Journal of the American College of Radiology. 2009;6(6):393–6.

[18] Yamada K, Slanetz PJ, Boiselle PM. Perceived Benefits of a Radiology Resident Mentoring Program: Comparison of Residents With Self-Selected vs Assigned Mentors. Canadian Association of Radiologists Journal. 2014 May 1;65(2):186–91.

